# Symptomatic chikungunya and chronic post-infection arthralgia in a highly endemic setting in Northeastern Brazil, 2018-2019: clinical characteristics, prevalence and associated factors

**DOI:** 10.1101/2025.06.29.25330524

**Authors:** Carolline A Mariz, Natália Menezes N de Oliveira, Sílvia Carla de A Alexandre, Isabelle Viana, Clarice N L de Morais, Ernesto T A Marques, Thomas Jaenisch, Wayner Vieira de Souza, Maria de Fátima P Militão de Albuquerque, Carlos A A de Brito, Cynthia Braga

**Author notes:** Corresponding author, (CAM). Author contributions Conceptualization: Carolline A Mariz, Wayner V Souza, Maria de Fatima P Militão de Albuquerque, Thomas Jaenisch, Ernesto T A Marques, Carlos A A Brito, Cynthia Braga. Data curation: Carolline A Mariz, Wayner V Souza, Cynthia Braga. Formal analysis: Carolline A Mariz, Wayner V Souza, Cynthia Braga. Funding acquisition: Cynthia Braga, Thomas Jaenisch, Wayner V Souza, Maria de Fatima P Militão de Albuquerque. Investigation: Carolline A Mariz, Natália Menezes N de Oliveira, Sílvia Carla de A Alexandre, Clarice N L Morais, Ernesto T A Marques, Isabelle F T Viana, Cynthia Braga. Methodology: Carolline A Mariz, Wayner V Souza, Carlos A A Brito, Cynthia Braga. Project administration: Carolline A Mariz, Cynthia Braga. Supervision: Carolline A Mariz, Clarice N L Morais, Isabelle F T Viana, Cynthia Braga. Validation: Carolline A Mariz, Wayner V Souza, Maria de Fatima P Militão de Albuquerque, Carlos A A Brito, Cynthia Braga. Writing – original draft: Carolline A Mariz, Wayner V Souza, Maria de Fatima P Militão de Albuquerque, Carlos A A Brito, Cynthia Braga. Writing – review & editing: Carolline A Mariz, Natália Menezes N de Oliveira, Sílvia Carla de A Alexandre, Isabelle Viana, Clarice N L de Morais, Ernesto T A Marques, Thomas Jaenisch, Wayner Vieira de Souza, Maria de Fátima P Militão de Albuquerque, Carlos A A de Brito, Cynthia Braga.

## Abstract

Chikungunya, an *Aedes*-borne disease, poses a significant global health threat due to its substantial morbidity burden. Symptomatic chikungunya virus (CHIKV) infection and persistent post-infection arthralgia, along with their risk factors, exhibit considerable geographic and interstudy variation. We estimated the prevalence of symptomatic cases and chronic arthralgia among CHIKV-exposed participants in a household-based survey conducted in a large northeastern Brazilian city approximately three years after the virus’s emergence (2018–2019). Sociodemographic and clinical data were collected via interviews, and venous blood samples were tested for arboviruses (dengue-DENV, CHIKV, Zika) through IgM/IgG ELISA. Arthralgia severity was assessed via Visual Analog Scale (VAS). Prevalence estimates (95% CI) were calculated, and adjusted prevalence ratios (aPRs) were estimated using Poisson regression with robust variance to identify risk factors. Of 760 CHIKV-exposed participants, 70% (95% CI: 66.7–73.2; n=532) were symptomatic (symptomatic-to-asymptomatic ratio: 2.16:1). Prior DENV infection was detected in 93.8% (n=713). Among those with arthralgia (n=499), over 70% reported severe pain (VAS ≥8). Independent risk factors for symptomatic CHIKV included older age (aPR=1.21 [36–50 years], 1.19 [51–65 years]), female sex (aPR=1.19; 95% CI: 1.08–1.32), prior DENV exposure (aPR=1.45; 95% CI: 1.03–2.04), and lower family income (aPR=1.37 [>2–4 wages], 1.34 [≤2 wages]). Chronic arthralgia (persisting >90 days) occurred in 36.5% (n=182) of cases and was associated with older age (aPR=4.60 [51–65 years]), female sex (aPR=1.70; 95% CI: 1.29– 2.25), and severe acute pain (aPR=2.91; 95% CI: 1.86–4.55). Lower income (≤2 wages) was protective against chronic arthralgia (aPR=0.67; 95% CI: 0.50– 0.90). The high burden of chikungunya and chronic arthralgia in this population underscores the need for targeted interventions, particularly for older adults, women, and low-income groups. The association between prior DENV infection and symptomatic CHIKV suggests potential cross-viral interactions in co-endemic settings.

## Introduction

Chikungunya, a mosquito-borne disease primarily transmitted by Aedes aegypti and Aedes albopictus, has emerged as a significant global health problem, particularly in Asian, African and the Americas [1, 2]. Between 2011 and 2020, an estimated 19 million cases were reported worldwide, with approximately 80% occurring in Latin America and the Caribbean region [2].

Infection by the chikungunya virus (CHIKV, family Togaviridae, genus Alphavirus) has a wide spectrum of manifestations, ranging from asymptomatic or mild cases to severe and debilitating conditions, including neurological disorders and chronic arthralgia [3]. The prevalence of symptomatic chikungunya varies significantly across regions and populations, with an estimated rate of 75% (range: 63%-84%) [4]. The acute phase of chikungunya is primarily characterized by high fever (>39°C), rash, headache, arthralgia and myalgia—symptoms that overlap with other arboviral infections, such as dengue [4].

Arthralgia is one of the most prominent symptoms of chikungunya, occurring in approximately 90% of cases. It is typically bilateral and symmetrical, predominantly affecting the joints of the hands, feet, knees and wrists [4, 5]. Persistent arthralgia— defined as joint pain lasting beyond three months after the acute phase—is a major complication, affecting 30-60% of infected individuals [6–8]. A systematic review of 67 studies estimated the prevalence of chronic arthralgia at 44% after three months, declining to 34% after six months and 32% at twelve months [4].

The frequency of symptomatic chikungunya and persistent arthralgia post-infection, as well as the factors associated with these events, has shown high geographic heterogeneity and variation between studies [4, 7]. Both events have been linked to a number of individual factors, including sociodemographic characteristics (e.g., advanced age and female sex) [9, 10], comorbidities (diabetes, hypertension, ischemic heart disease, pre-existing joint disease), genetic polymorphisms and immunological factors [7, 11–13]. Viral strain-related factors, such as the CHIKV lineage, have also been associated with chronic arthralgia. Specifically, the Asian lineage has been associated with a higher incidence of inapparent chikungunya infections [14], whereas the Indian Ocean lineage (a sublineage of the East/Central/South African [ECSA] strain) has been linked to chronic arthralgia [15].

Since its first detection in Brazil in 2014, CHIKV has caused severe recurrent epidemics nationwide. From 2013 to 2022, seven epidemic waves were recorded, with the Northeast region—accounting for ∼65% of confirmed cases—being among the most affected [16]. We conducted a population-based seroprevalence survey in Recife, a major urban center in Northeastern Brazil, two years after the city’s first major CHIKV epidemic (2016) to evaluate the long-term transmission patterns of CHIKV and other arboviruses [17]. The results showed an overall CHIKV, DENV and Zika (ZIKV) seroprevalence of 35.7%, 88.7% and 37.2%, respectively. In this study, we estimated the prevalence of symptomatic CHIKV infection and persistent post-infection arthralgia, as well as investigated associated risk factors among the CHIKV-exposed participants in this survey.

## Material and methods

### Design, setting and study population

The multistage stratified cluster sampling population survey was conducted between August 2018 and February 2019. Details on the sampling methods and selection of study participants have been published elsewhere Recife, the capital of Pernambuco state, has a population of approximately 1.5 million inhabitants and one of Brazil’s highest population densities (7,000 inhabitants/km²) [18]. Since the first dengue epidemic in the 1980s, the city has experienced recurrent arbovirus outbreaks. Based on our seroprevalence estimate (35.7%, n=2.070), we project that approximately 600.000 residents were exposed to CHIKV between the 2015 emergence and 2018/2019 survey period [17].

All survey participants with positive serology for CHIKV were included in the study (IgM/IgG; n=760). The data analysis of chronic arthralgia was restricted to symptomatic cases (n=532).

### Data collection

In the serosurvey, we invited all eligible residents of selected households within the study’s age range to participate. Participants read and signed the Informed Consent Form before study staff collected sociodemographic and clinical data through interviews using a structured questionnaire. For participants with arthralgia, pain intensity was assessed using a visual analog scale (VAS; 0 = no pain, 10 = very severe pain) [18]. Subsequently, a venous blood sample was collected for serological testing. Further detail on data collection was previously described [17].

### Laboratory procedures

Detection of anti-CHIKV IgG and IgM antibodies was performed using commercial ELISA kits (Euroimmun, Lubeck, Germany) and the results were interpreted according to the manufacturer’s instructions (CHIKV IgG or IgM absorbance at 450 nm/calibrator ratios were considered negative at <0.8, indeterminate at ≥0.8 to <1.1, and positive at ≥1.1). Previous or recent exposure to ZIKV was assessed by detecting IgG ELISA (Euroimmun, Lubeck, Germany) and by detecting IgG3 antibodies against the ZIKV NS1 protein through an in-house ELISA [17]. Anti-DENV IgG was determined by an in-house indirect ELISA method, as described elsewhere [19]. Results were interpreted as positive when the sample absorbance at 450 nm/calibrator ratio ≥3.62.

### Case definitions

Symptomatic chikungunya: defined as laboratory detection of anti-CHIKV antibodies (IgG and/or IgM) accompanied by one of the following conditions:

(1) A self-reported history of chikungunya within the preceding three years; or
(2) No self-reported history of chikungunya but self-reported dengue or Zika in the previous three years, with negative serology for ZIKV (anti-ZIKV IgG, IgM, and/or IgG3). (IgG, IgM, and/or IgG3).

This second case definition criterium was established due the significant clinical overlap between arboviral infections (dengue, Zika, and chikungunya); the recent and near-simultaneous introduction of ZIKV and CHIKV in the study region [20]; and evidence of low DENV circulation during survey data collection [20, 21].

Chronic arthralgia: defined as the report of joint pain persisting for >90 days after the acute phase of infection.

### Exposed variables

Demographic: Age group (5–19, 20–35, 36–50, and 51–65 years), sex, self-reported race/skin color (Brown, Black, White, and Asian), educational level (elementary, high school, university/postgraduate), marital status (with or without partner), head of family income in minimum wages (≤2, >2 to ≤4, and >4).

Clinical: Comorbidities (hypertension, diabetes, chronic heart disease, nephropathy, or others), DENV immune status (anti-DENV IgG), joint pain intensity (assessed by visual analogue scale, VAS [1-10 score]), severity of joint pain (very severe=VAS>8 ), self-reported chikungunya symptoms (fever [and duration], headache, rash, conjunctivitis, photophobia, myalgia, eye pain, arthralgia, edema, anorexia, prostration, bleeding, limb paralysis, meningitis, pruritus, diarrhea, and nausea/vomiting), as well as joints affected by pain.

### Data analysis

Data management was performed using the REDCap electronic platform (hosted at the University of Heidelberg, Germany), and statistical analyses were conducted using Stata software (version 15; StataCorp, College Station, TX, USA).

We first characterized the study population by describing the frequency distributions of key variables, presenting means ± standard deviations for continuous variables and proportions for categorical variables. Difference between means were assessed by Kruskal Wallis (KW) test (p-value<0.05).

Prevalence estimates with 95% confidence intervals (95% CI) were calculated for symptomatic chikungunya and persistent arthralgia.

To assess associations between exposure variables and the clinical outcomes (chikungunya or chronic arthralgia), we estimated both crude (PR) and adjusted prevalence ratios (aPRs) using Poisson regression with robust variance. This method is preferable for estimating association measures for high-prevalence outcomes [22]. Variables associated with the outcomes at p < 0.20 in univariable analyses were included in the multivariable regression model.

Initially, we examined the crude associations of sociodemographic variables (age group, sex, self-reported race/skin color, educational level, marital status, head of family income) and clinical variables (Comorbidities, DENV-immune status, severity of joint pain) with the outcomes analyzed. The severity of joint pain was included only in the analysis of persistent arthralgia. Variables significantly associated with the outcome (p<0.05) were retained in the final multivariable model.

### Ethical issues

This study was approved by the Research Ethics Committee of the Aggeu Magalhães Institute, Pernambuco, Brazil (CAEE n° 79605717.9.0000.5190; approval number: 2.734.481). All participants read and signed the Informed Consent Form as required by the ethics requirements of Brazilian National Health Council Resolution N°466/12. Participants aged 5 to 18 years provided oral and written assent.

## Results

A total of 760 survey participants were exposed to CHIKV. The median age was 38 years (range: 5–65), with a predominance of females (60.3%, 458/760), 54% (411/760) self-identifying as mixed-race (brown skin color) and 30.7% (175/760) reporting comorbidities. Prior DENV infection, determined by anti-DENV IgG seropositivity, was detected in 93.8% (n = 713/760) of the study population.

Of these, 532 met the symptomatic case definition criteria, resulting in a symptomatic chikungunya prevalence of 70% (95% CI: 66.7–73.2) and a symptomatic-to-asymptomatic ratio (SAR) of 2.16:1 (95% CI: 1.89:1–2.37:1).

The most frequently reported symptoms among symptomatic cases were arthralgia (93.8%), myalgia (91.3%), fever (89.8%), prostration (89.8%), and headache (81.4%). Rash, photophobia, eye pain, and edema were reported in approximately half of cases (Table 1).

**Table 1.** Frequency distribution of the reported symptoms among cases of symptomatic chikungunya. Recife, Recife, Northeast Brazil, 2018-2019 Characteristics/symptoms n = 532.

| Characteristics/symptoms | n = 532 |
| --- | --- |
| Arthralgia | 499 (93.8) |
| Myalgia | 486 (91.3) |
| Severe/very severe myalgia, n (%) | 341 (70.6) |
| Fever, n (%) | 478 (89.8) |
| Duration of fever (in days), mean (Standard deviation, SD) | 4.7 (4) |
| Prostration, n (%) | 478 (89.8) |
| Headache, n (%) | 433 (81.4) |
| Anorexia, n (%) | 421 (79.1) |
| Severe/very severe headache (VAS* >8) | 221 (51.2) |
| Rash, n (%) | 293 (55.1) |
| Photophobia, n (%) | 282 (53.0) |
| Eye pain, n (%) | 298 (56.0) |
| Conjunctivitis, n (%) | 200 (37.6) |
| Edema, n (%) | 246 (46.3) |
| Bleeding, n (%) | 4 (0.7) |
| Paralysis of limbs, n (%) | 16 (3.0) |
| Meningitis, n (%) | 2 (0.4) |
| Itching, n (%) | 97 (18.2) |
| Diarrhea, n (%) | 35 (4.6) |
| Nausea/vomit, n (%) | 99 (13.0) |
| **Characteristics of arthralgia** | n=499 |
| Intensity of pain, mean (SD) (VAS*) | 8.3 (1.9) |
| Severe/very severe arthralgia, n (%) (VAS ≥ 8) | 361 (72.3) |
| **Joints affected, n (%)** |  |
| Hands | 429 (89.9) |
| Wrist | 431 (86.4) |
| Foot | 424 (84.9) |
| Ankles | 419 (83.9) |
| Knees | 406 (81.4) |
| Elbows | 312 (62.5) |
| Shoulders | 323 (64.7) |

\* Visual analog scale

Among participants reporting arthralgia (n=499), over 70% experienced severe pain (VAS score ≥ 8), with a mean pain intensity of 8.3 ± 1.9 (median 9: range 1-10). The most frequently affected joints were those of the hands (89.9%), wrists (86.4%), feet (84.9%), ankles (83.9%), and knees (81.4%). Elbows and shoulders were involved in over half of cases (Table 1).

Pain intensity was significantly higher in those with arthralgia persisting >90 days (mean ± SD: 9.1 ± 1.5) compared to those with shorter symptom duration (7.9 ± 2.0; KW χ² = 54.61, *p* < 0.001). Similarly, females reported higher pain intensity than males (8.7 ± 0.1 vs. 7.6 ± 0.2; KW χ² = 30.37, p < 0.001), as did participants with comorbidities compared to those without (8.6 ± 0.15 vs. 8.2 ± 0.11; KW χ² = 6.71, p = 0.009).

The univariable regression analysis showed that age group, sex, marital status, head of household income, education level, comorbidities, and prior DENV exposure (IgG anti-DENV) were statistically (p < 0.20) associated with symptomatic chikungunya and these variables were retained for multivariable analysis (Table 2). In the adjusted model, older age (≥36 years; aPR = 1.21 for 36–50 years and 1.19 for 51–65 years), female sex (aPR = 1.19; 95% CI: 1.08-1.32) and prior DENV exposure (aPR = 1.45; 95% CI: 1.03–2.04) were independently associated with an increased risk for symptomatic chikungunya. Conversely, higher household income (>4 minimum wages; aPR = 1.00 [reference]) was protective compared to lower income families (aPR = 1.37 for >2–4 wages; aPR = 1.34 for ≤2 wages) (Table 2).

**Table 2.**
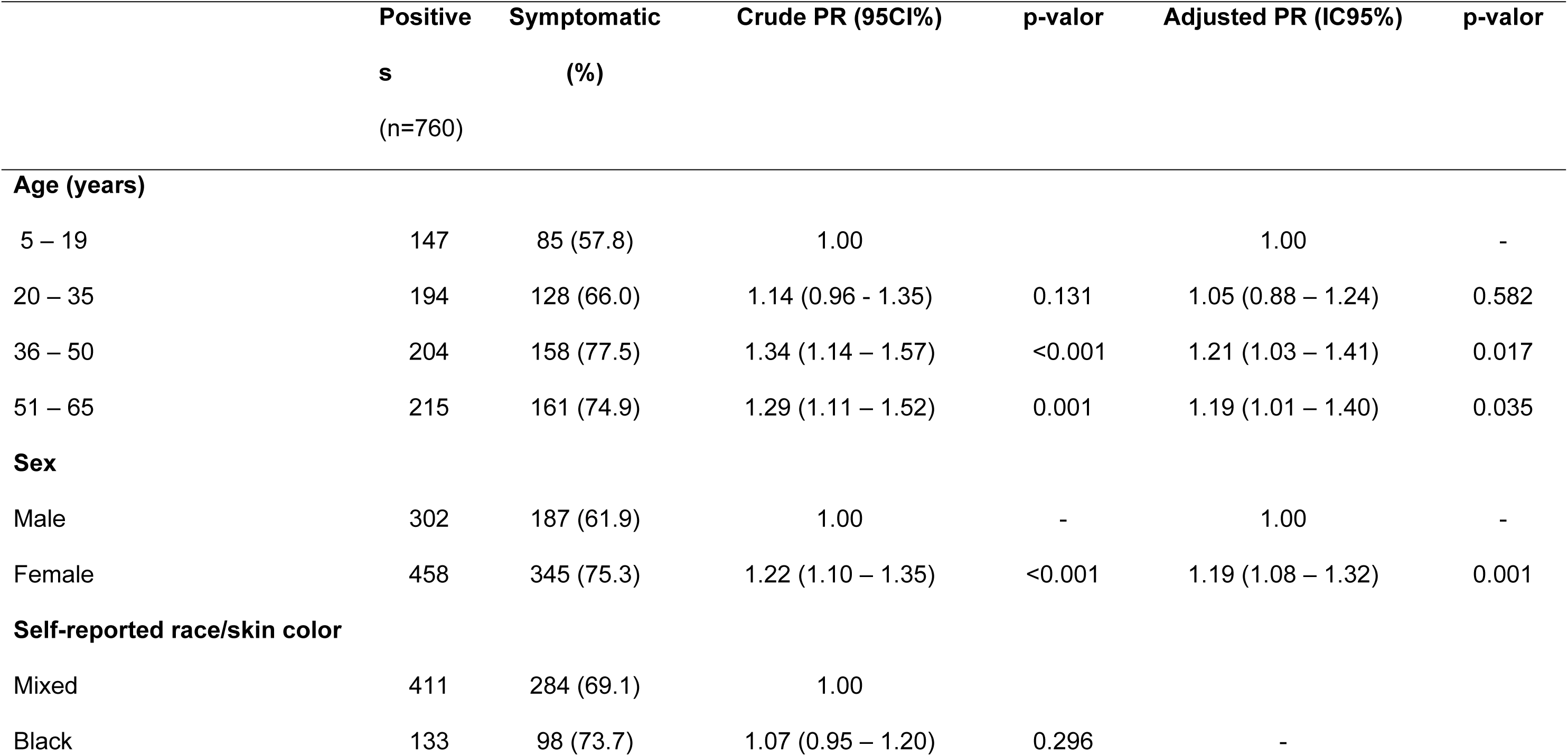
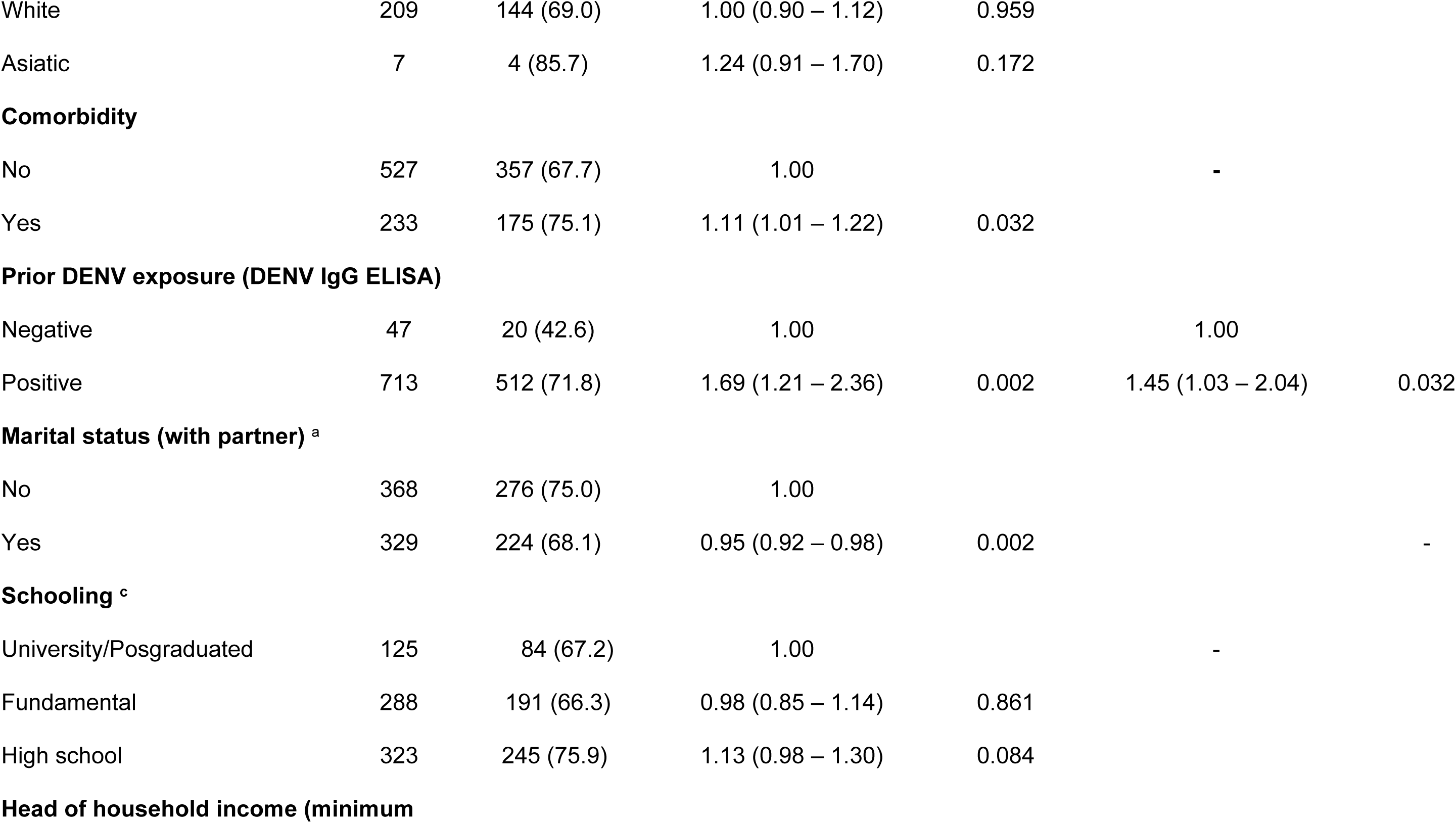
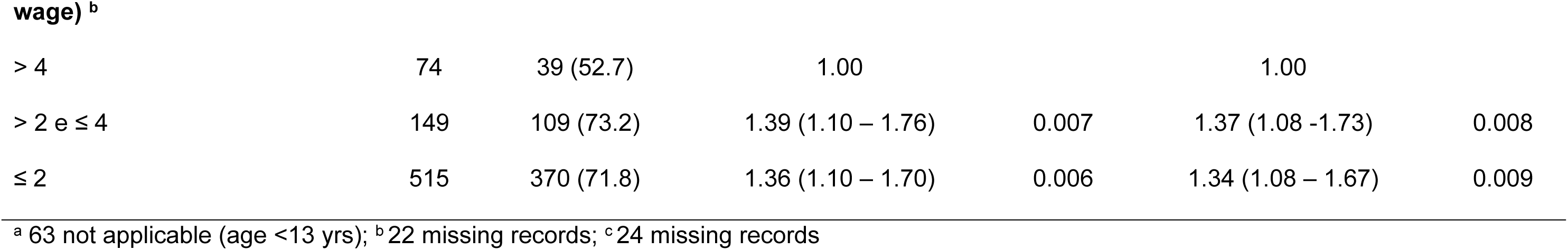
Crude and adjusted prevalence rates (PR) of the association of sociodemographic and clinical characteristics with symptomatic chikungunya. Recife, Northeast Brazil, 2018-2019.

We next examined factors associated with chronic arthralgia (>90 days) among symptomatic cases. Among symptomatic cases with arthralgia (n=499), 36.5% (95% CI: 32.4–40.8; n=182) had persistent symptoms (>90 days), classified as chronic arthralgia. Univariate logistic regression identified age, sex, head of the household income, comorbidities and severe acute joint pain (VAS ≥8) as factors statistically associated with chronic arthralgia (p<0.20). These variables were included in the multivariable regression model. In the adjusted model, older age (≥20 years; demonstrating a dose-response association: aPR = 2.39 for 20–35 years, aPR = 4.10 for 36–50 years, and aPR = 4.60 for 51–65 years), female sex (aPR = 1.70; 95% CI: 1.29–2.25), and severe acute joint pain (aPR = 2.91; 95% CI: 1.86–4.55) were independently associated with an increased risk of chronic arthralgia (Table 3). Comorbidities (e.g., hypertension/diabetes) were associated with chronic arthralgia in unadjusted analysis (PR = 1.64, p <0.01) but not in the adjusted model. The strongest association was in the 51– 65-year age group (aPR = 4.60; 95% CI: 2.10 – 10.03), with a 4.6-fold higher prevalence of chronic arthralgia than the reference group (5–19 years). Participants living in households with lower income (≤2 minimum wages) had approximately 33% lower prevalence of chronic arthralgia (aPR=0.67; 95% CI: 0.50–0.90) compared to the highest income group (>4 wages) (Table 3).

**Table 3.**
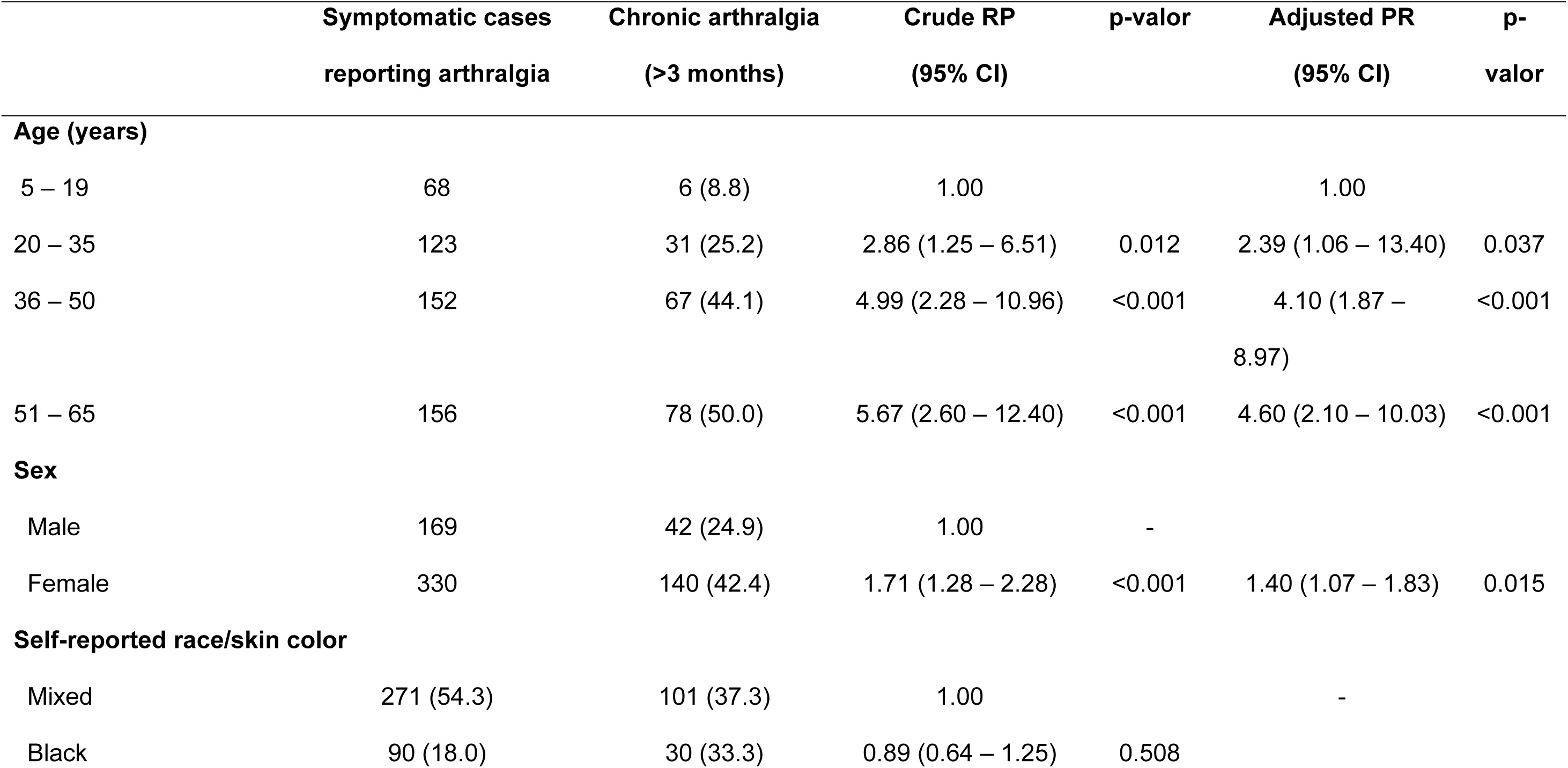
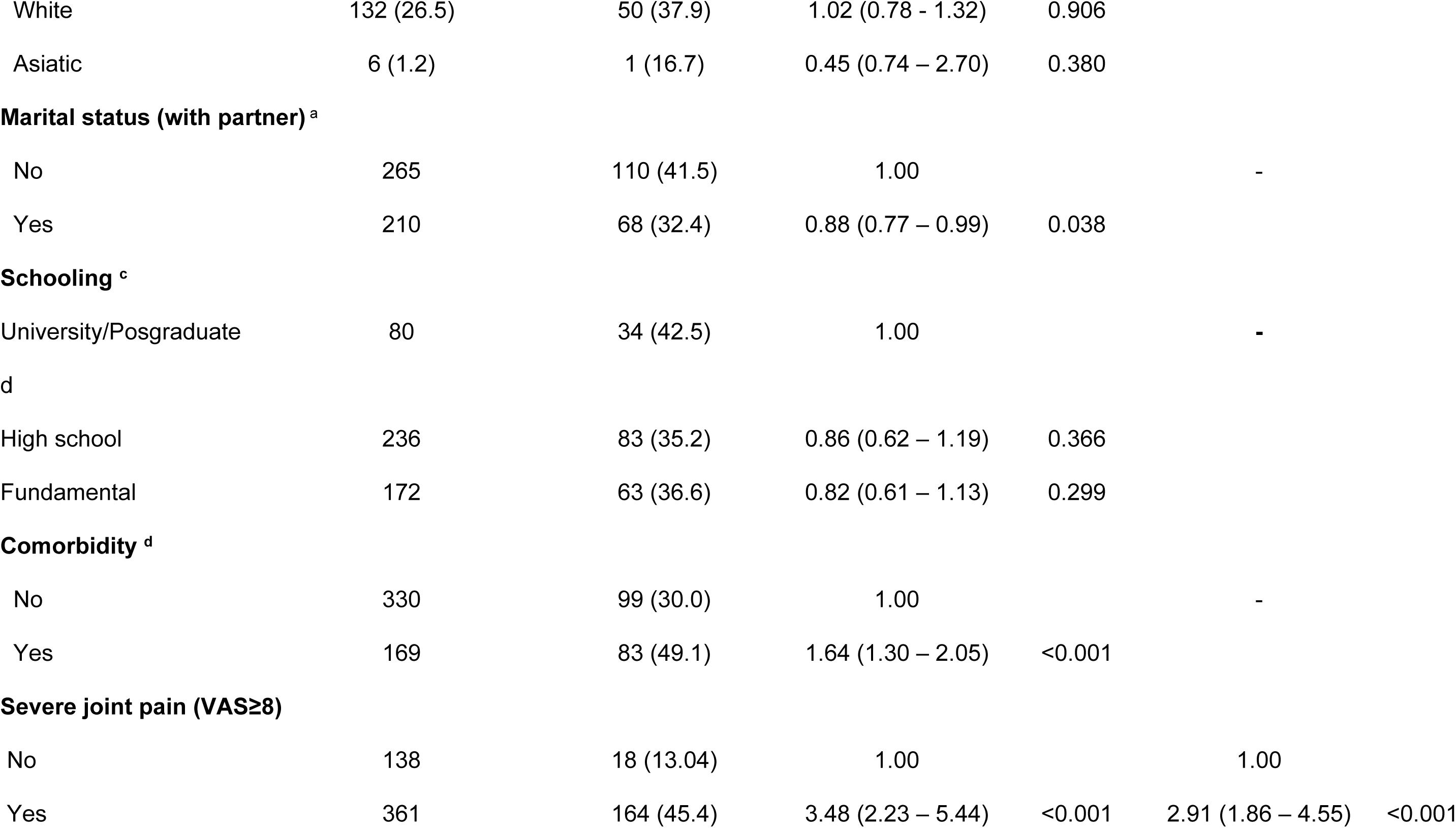
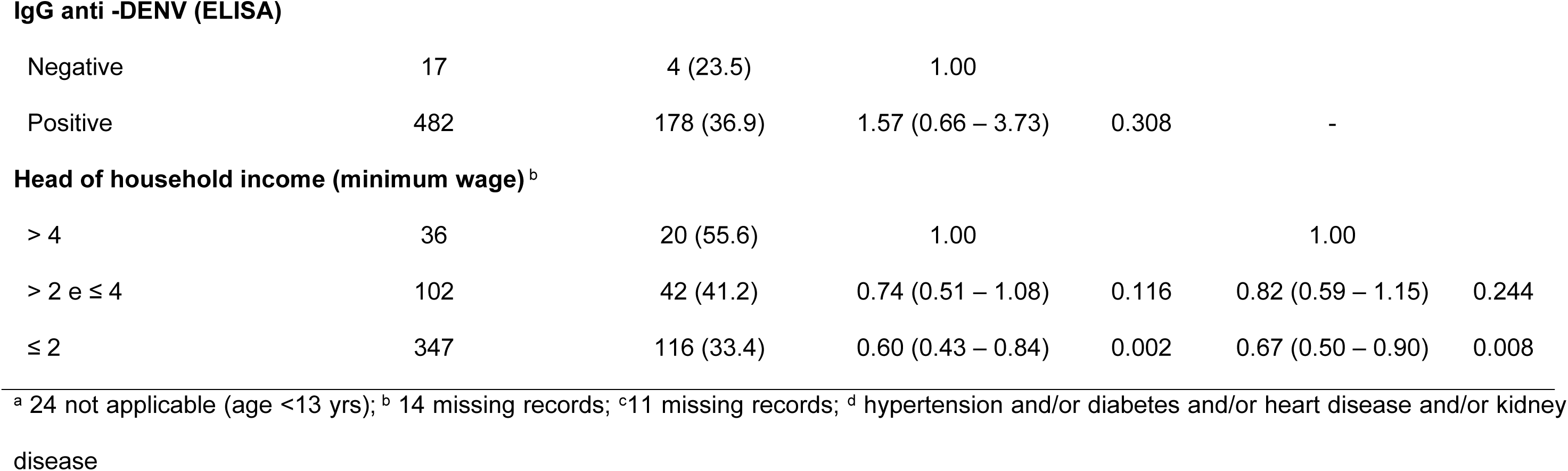
Crude and adjusted prevalence rates (PR) of the association of sociodemographic and clinical characteristics with chronic arthralgia among symptomatic cases of chikungunya (n=499). Recife, Northeast Brazil, 2018-2019.

## Discussion

In this study, we estimated the prevalence and analyzed factors associated with symptomatic chikungunya and chronic arthralgia in a population living in an arbovirus-endemic area, where approximately 90% had prior DENV exposure. Consistent with previous studies [4, 23], our findings highlight the substantial burden of chikungunya in the study setting, showing a 70% prevalence of symptomatic cases (SAR = 2.16:1). Moreover, 40% of participants reporting acute-phase arthralgia progressed to chronic arthralgia (persisting >90 days).

The high prevalence of previous DENV infection, together with the observed associations between demographic factors and clinical outcomes (symptomatic CHIKV infection and chronic arthralgia), raise some questions about host susceptibility and potential interactions between these arboviruses.

In line with literature data [4, 24], the most frequently reported symptoms among symptomatic cases were arthralgia (93.8%), myalgia (91.3%), fever (89.8%), and prostration (89.8%). The arthralgia predominantly affected small joints— particularly the hands, wrists, and feet—aligning with findings from other studies [13].

Notably, over 70% of individuals who reported arthralgia during the acute phase had severe pain, with a mean pain intensity of 8.3 ± 1.9 on the VAS. Pain scores were significantly higher in females, among those who developed chronic arthralgia, and in individuals with comorbidities—factors previously associated with prolonged musculoskeletal symptoms [7, 25].

Few studies have assessed joint pain intensity following CHIKV infection using the VAS [25–27]. These studies reported pain intensity as moderate to severe, highlighting the clinical significance of the disease. However, our findings demonstrated even higher pain scores than those previously documented. Two cross-sectional studies—one conducted on Réunion Island (mean VAS = 5.8 ± 2.1) [26] and another in northeastern Brazil (mean VAS = 6.5 ± 2.0) [27] — reported lower pain intensity than observed in our study population. In contrast, a prospective study of acutely ill patients admitted to hospitals in Mexico reported higher mean pain scores than those in our study [25].

The prevalence of symptomatic chikungunya (70%) in our study population was high, aligning with rates reported in other regions [4,23]. Globally, studies have documented substantial variability in symptomatic chikungunya prevalence [4, 9, 14, 23, 28, 29], suggesting that host- and virus-specific factors may shape local epidemiological patterns. Despite this variability, only three studies have analyzed risk factors, and their findings remain inconsistent [9, 14, 23], highlighting a critical evidence gap.

The multivariable regression analysis of the associated factors with symptomatic chikungunya showed that older age (>36 years), female sex, head of household income, and prior DENV exposure were independently (p < 0.05) associated with symptomatic chikungunya.

The increase in the prevalence of symptomatic chikungunya with age is in line with findings of a large prospective study conducted during a chikungunya epidemic in Nicaragua, which reported a linear increase in the probability of symptomatic infection with age [14]. Similarly, a post-epidemic serosurvey in two cities in northeastern Brazil supported this association [28]. The higher prevalence of symptomatic chikungunya among those with older age is explained by immunosenescence, comorbidities, and cumulative viral past exposures. Age-related dysregulation of cellular immunity may delay viral clearance and prolong inflammatory responses, exacerbating joint damage and persistent arthralgia [30, 31]. Additionally, comorbidities such as diabetes, hypertension, and cardiovascular disease —more prevalent in older populations—may worsen outcomes by impairing innate immune responses [32].

However, our results contrast with a population-based survey in Malaysia, where older individuals (≥58 years) was protective to symptomatic infection compared to younger adults (18–36 years) [9]. Another survey in the U.S. Virgin Islands found no significant age-related differences in symptomatic disease or long-term joint pain [23]. These discrepancies may arise from variations in host genetic profiles [33], circulating viral lineages [14], or cultural differences in pain perception [34].

Similar to the observed differences in pain intensity between sexes, the prevalence of symptomatic chikungunya in our study was approximately 20% higher in females than males (aPR 1.19). These findings align with studies reporting a higher frequency and severity of acute symptoms among females during CHIKV epidemics [35] [7, 25] and, may be attributed to hormonal influences, particularly estrogen and progesterone, which modulate immune responses and can exacerbate inflammatory reactions to infection [36]. Additionally, sex-based differences in pain perception may contribute to these findings, as multiple studies suggest that females experience greater clinical pain sensitivity and heightened responses to experimentally induced pain compared to males [37]. However, our result contrast with other studies that found no sex-based differences in symptomatic infection rates [23,14].

Surprisingly, prior DENV infection (93.8% seropositivity in the study population) was significantly associated with symptomatic CHIKV infection (aPR = 1.45; 95% CI: 1.03–2.04), suggesting a potential interaction between DENV and CHIKV infections in co-endemic settings that may exacerbate clinical outcomes. This epidemiological observation aligns with in vitro evidence demonstrating that prior DENV infection enhanced CHIKV replication [38]. Specifically, subneutralizing levels of anti-DENV antibodies were shown to enhance CHIKV infection in human cells (K562 cells), suggesting a possible cross-reactive antibody-dependent enhancement (ADE) mechanism [38]. Another experimental study further supports that ADE may contribute to CHIKV disease severity [29] . However, large-scale clinical or epidemiological studies are needed to confirm this association and assess its public health implications in co-endemic regions.

The multivariable analysis further identified lower income (≤4 minimum wages) as an independent risk factor for symptomatic CHIKV infection. Notably, households earning less than two minimum wages showed significantly higher prevalence of symptomatic cases. This association may reflect reduced healthcare-seeking behavior due to systemic barriers in healthcare access. While several studies have established links between socioeconomic status and Aedes-borne disease incidence [39, 40], research specifically examining socioeconomic factors in relation to symptomatic chikungunya remains limited. Our findings are partially consistent with a Malaysian serological survey that identified higher education levels as a protective factor for symptomatic infection[9] .

Approximately 40% of individuals reporting arthralgia developed chronic symptoms, a finding consistent with prevalence rates reported in systematic reviews and meta-analyses of CHIKV clinical outcomes in endemic regions [4, 7, 41], highlighting the significant long-term morbidity associated with CHIKV infection.

In the final multivariable model, older age (>20 years; dose-dependent effect), female sex, severe acute joint pain and low income of the head of household (≤2 minimum wages) demonstrated strong associations with chronic arthralgia. While comorbidities showed a strong association with chronic arthralgia in univariate analysis, this relationship disappeared in the multivariate model, suggesting possibly competing risk factors in our population. The age-dependent increase in chronic arthralgia aligns with findings from multiple studies on symptomatic chikungunya [6, 7, 34]. This trend may reflect age-related immune dysregulation, leading to impaired viral clearance and prolonged inflammation [31], or joint vulnerability due to degenerative changes [42] .

The prevalence of chronic arthralgia, as observed in the analysis of factors associated with symptomatic chikungunya, was higher in females (aPR = 1.40) than in males in our study population, and this association has been reported in several studies [25, 43]. The greater occurrence of symptoms and severity of symptoms in females has been explained by immune-related genes, caused by biased responses from the X chromosome, which harbors the majority of immune-related genes, or by the influence of hormonal factors [36, 44]. Female sex hormones (estrogen and progesterone) may modulate immune responses, leading to stronger inflammatory reactions [44], which may worsen joint pain and swelling. In addition to biological mechanisms, psychosocial factors could also play a role in these sex differences. Women would report symptoms more frequently and exhibit greater healthcare-seeking behavior, leading to apparently higher severity in studies [37, 45].

The observed association between severe joint pain and chronic arthralgia aligns with findings from a retrospective cohort study of 147 CHIKV-infected patients in Réunion Island [46], which reported a five-fold higher prevalence of persistent symptoms among individuals who experienced severe pain (assessed via a numerical rating scale [NRS]) during the acute phase of infection compared to those with milder pain. These findings reinforce evidence that greater pain intensity during the acute phase—potentially indicative of an exacerbated inflammatory immune response and viral persistence in musculoskeletal tissues [12] — serves as a predictor of chronic joint symptoms, highlighting the need for targeted clinical monitoring in such cases.

Interestingly, our analysis found an inverse association between household income and chronic arthralgia, with lower-income households (≤2 minimum wages; aPR = 0.67 [95% CI: 0.50 – 0.90]) exhibiting reduced risk compared to higher-income counterparts (>4 MW). This aligns with prior reports of higher chronic arthralgia prevalence in affluent populations, such as French workers in Réunion Island, relative to lower-income groups in India[7]. A subsequent systematic review of 38 global studies further supported this trend, noting elevated chronic chikungunya prevalence in high-income countries [41], though confidence intervals overlapped. The findings can be explained by sociocultural factors that influence the perception of pain in these populations. It is assumed that chronic pain and symptoms can be normalized, leading individuals to tolerate pain and consider it expected. Although studies indicate an inverse relationship between socioeconomic status and chronic pain—with individuals of lower socioeconomic status tending to experience greater pain chronicity [47] —evidence suggests that social exclusion correlates with reduced pain sensitivity, often described as ’numbing’ [48] .

In this study, clinical outcome data were obtained through self-reported symptoms and pain scores, rendering them susceptible to information bias, which may compromise the validity of the results. Additional limitations include the cross-sectional design, the lack of biomarker measurements or other clinical exams, and potential unmeasured confounding factors—all of which hinder causal inference. Nevertheless, our study was conducted shortly after the emergence of CHIKV and its subsequent epidemic in the study setting [17] , with evidence of low DENV circulation during the study period [20, 21]. These factors likely reduced the risk of information bias and cross-diagnosis between dengue and chikungunya. Strengths of the study include a large population sample with laboratory-confirmed CHIKV, ZIKV, and DENV exposure, detailed symptom characterization (e.g., VAS, joint-specific data), and the use of Poisson regression — the most appropriate method for estimating prevalence ratios (PRs) in high-frequency event analyses [22].

In summary, our study provides evidence for the high prevalence of symptomatic chikungunya and chronic arthralgia in a population previously exposed to DENV, with age, sex, prior DENV infection, and socioeconomic status emerging as critical determinants of clinical outcomes. The observed associations—particularly the potential role of DENV-CHIKV immunologic interactions and sociodemographic disparities in pain severity and chronicity— warrant further investigation through longitudinal studies incorporating biomarkers and viral genomic data.

Despite the inherent limitations of the cross-sectional study design, our findings corroborate the evidence on the long-term morbidity of chikungunya in this context and highlight the need for targeted interventions in co-endemic settings, especially targeting higher-risk groups such as the elderly, women, and low-income populations. Future research should prioritize mechanistic studies to clarify the interplay between arboviral infections and host factors, as well as community-based strategies to mitigate chronic disability associated with CHIKV.

## Data Availability

All relevant data are within the manuscript

